# Modelling SARS-CoV-2 Dynamics: Implications for Therapy

**DOI:** 10.1101/2020.03.23.20040493

**Authors:** Kwang Su Kim, Keisuke Ejima, Yusuke Ito, Shoya Iwanami, Hirofumi Ohashi, Yoshiki Koizumi, Yusuke Asai, Shinji Nakaoka, Koichi Watashi, Robin N. Thompson, Shingo Iwami

**Author notes:** To whom correspondence may be addressed. (R.N.T.) and (S.I.).

## Abstract

The scientific community is focussed on developing antiviral therapies to mitigate the impacts of the ongoing novel coronavirus disease (COVID-19) outbreak. This will be facilitated by improved understanding of viral dynamics within infected hosts. Here, using a mathematical model in combination with published viral load data collected from the same specimen (throat / nasal swabs or nasopharyngeal / sputum / tracheal aspirate), we compare within-host dynamics for patients infected in the current outbreak with analogous dynamics for MERS-CoV and SARS-CoV infections. Our quantitative analyses revealed that SARS-CoV-2 infection dynamics are more severe than those for mild cases of MERS-CoV, but are similar to severe cases, and that the viral dynamics of SARS-CoV infection are similar to those of MERS-CoV in mild cases but not in severe case. Consequently, SARS-CoV-2 generates infection dynamics that are more severe than SARS-CoV. Furthermore, we used our viral dynamics model to predict the effectiveness of unlicensed drugs that have different methods of action. The effectiveness was measured by AUC of viral load. Our results indicated that therapies that block *de novo* infections or virus production are most likely to be effective if initiated before the peak viral load (which occurs around three days after symptom onset on average), but therapies that promote cytotoxicity are likely to have only limited effects. Our unique mathematical approach provides insights into the pathogenesis of SARS-CoV-2 in humans, which are useful for development of antiviral therapies.

**Significance Statement:** Antiviral agents with different mechanisms of action have different curative effects depending on precisely when therapy is initiated. Based on a model of viral dynamics, parameterised using viral load data from SARS-CoV-2 infected patients reported by Zou *et al. (1)*, computer simulations were performed. We propose that effective treatment of SARS-CoV-2 infection requires an appropriate choice of class-specific drugs and initiation timing as reported for treatment of other viral infections (2); otherwise, antivirals do not have a significant effect on the within-host viral dynamics of SARS-CoV-2 and are wasted.

---

The ongoing coronavirus disease 2019 (COVID-19) outbreak was first reported in Wuhan, China in late December 2019 (3, 4). Since then, the causative agent (severe acute respiratory syndrome coronavirus 2, SARS-CoV-2) has been transmitted elsewhere in China and to 80 other countries and territories around the world. The number of confirmed cases currently stands at 139,061 (as of 13 March 2020). The possibility of presymptomatic or asymptomatic cases (5), combined with underreporting of symptomatic infections, suggests that the true number of cases is likely to be even higher than this.

Antiviral drugs (for treatment and to avoid onwards transmission) and a vaccine (for prevention) are currently under development to counter this outbreak. To aid the development process, characterisation of the viral dynamics of SARS-CoV-2 both *in vivo* and *in vitro* is crucial. The virus has been isolated, genome sequencing has been completed and the resulting data were made publicly available early in the outbreak (6, 7). Furthermore, the viral load in upper respiratory specimens (throat and nasal swabs) of infected patients over 20 days after symptom onset has been reported (2). However, the viral dynamics of SARS-CoV-2 infections have not been studied quantitatively, and the data have not been compared with analogous datasets for other coronaviruses. Such quantitative analyses are informative for the development of antiviral agents, addressing questions such as the optimal viral-host processes for antiviral drugs or vaccines to target.

## Results and Discussion

### Characterising SARS-CoV-2 and MERS-CoV infections by analysing viral load measurements collected from throat swabs

We analysed data describing SARS-CoV-2 viral loads reported by Zou *et al. (1)* and MERS-CoV viral loads reported by Oh *et al*. (8) using a simple mathematical model (see **Methods**). To consider inter-individual variations in viral loads, a nonlinear mixed-effect modelling approach was employed to estimate parameters (see **Methods**). The estimated parameters and initial values are listed in **Table 1**, and the typical behaviour of the model using these best-fit parameter estimates is shown together with the data in **Fig. 1A** for SARS-CoV-2 (pink) and MERS-CoV (black and grey for severe and mild case, respectively). In addition, to parameterise and compare these coronaviruses infections, we calculated the following important quantities (**Fig. 2**) using estimated parameter values; the mean length of virus production of an infected cell (*L* = 1/*δ*), the within-host basic reproduction number (*R*_0_ = *γ*/*δ*) which is the average number of newly infected cells produced by any single infected cell (9), and the critical inhibition rate (*C*^*∗*^ = 1 − 1/*R*_0_) induced by antivirals to prevent primary virus infection (10, 11). We showed that *L* is not significantly different for SARS-CoV-2 and MERS-CoV. However, interestingly, we found that *R*_0_ and *C*^*∗*^ for SARS-CoV-2 are significantly different from analogous values for mild cases of MERS-CoV (*p* = 8.9 × 10^−4^ and 2.0 × 10^−6^ by the bootstrap *t*-test, respectively), but not for severe MERS-CoV (*p* = 0.41 and 0.41) (see **Fig. 2**), although we were unable to separate mild and severe cases of SARS-CoV-2 due to limited clinical information for the cases. This demonstrates that SARS-CoV-2 causes infection more effectively than in mild cases of MERS-CoV, but a general SARS-CoV-2 infection follows infection dynamics that are similar to severe cases due to MERS-CoV. In addition, as a median estimate, 65% inhibition of the initial virus expansion is required to prevent the establishment of SARS-CoV-2 infection (we provide a detailed analysis later). We also calculated the duration of infection in which the viral load is above the detection limit (*T*_*V*_) in **Table 1**, showing that SARS-CoV-2 is maintained in hosts for more than a week based on the median estimate.

**Fig. 1.**
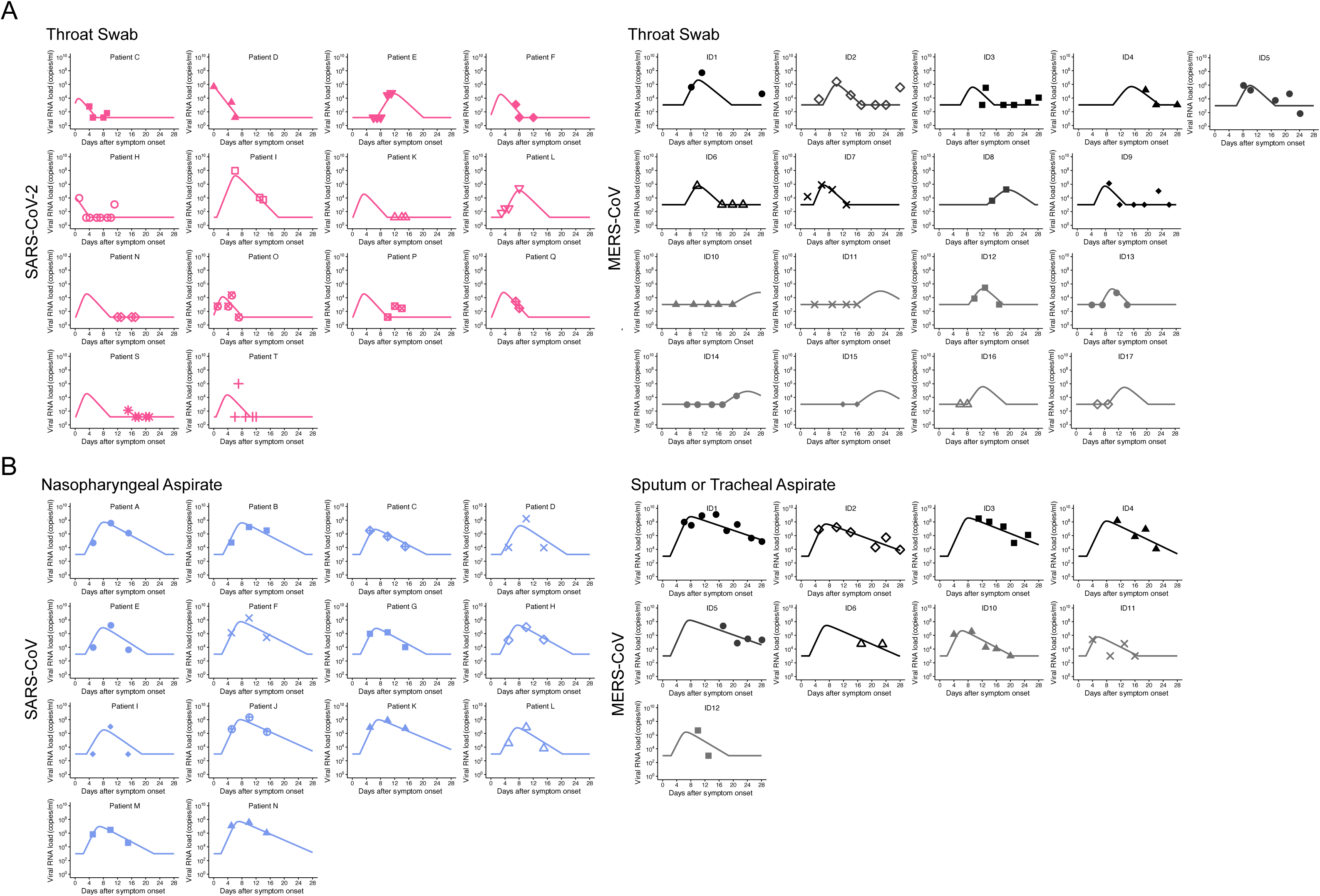
Mathematical model outputs for individual patients based on fits to viral load data for SARS-CoV-2, MERS-CoV and SARS-CoV. Viral loads were measured using throat swabs for SARS-CoV-2 and MERS-CoV (**A**) and nasoparyngeal/sputum swabs or tracheal aspirate for SARS-CoV and MERS-CoV (**B**) infected patients. Severe and mild cases of MERS-CoV are shown in black and gray, respectively. Note that the detection limits of measurements of SARS-CoV-2, MERS-CoV and SARS-CoV viral loads are 14.6, 1000 and 1000 copies/ml, respectively.

**Fig. 2.**
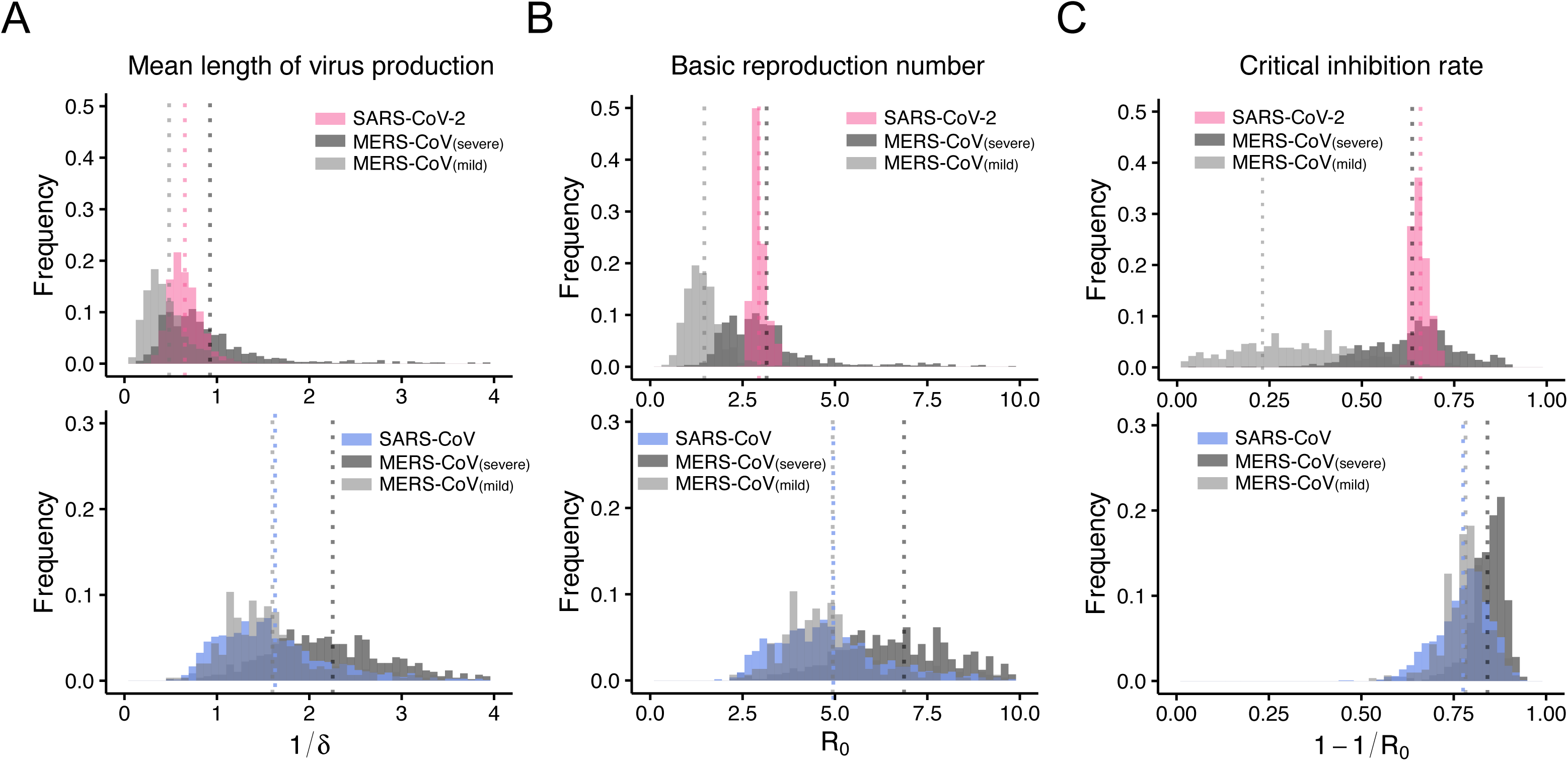
Characterisation and comparison of SARS-CoV-2, MERS-CoV and SARS-CoV infection dynamics *in vivo*. Distribution of estimates for (**A**) the mean duration of virus production of an infected cell, *L* = 1/*δ*, (**B**) the within-host basic reproduction number, *R*_0_ = *γ*/*δ*, and (**C**) the critical inhibition rate, *C*^*∗*^ = 1 − 1/*R*_0_. Estimates of *R*_0_ and *C*^*∗*^ for SARS-CoV-2 are significantly different compared to analogous estimates for mild cases of MERS-CoV, but not compared to estimates for severe MERS-CoV. On the other hand, estimates for SARS-CoV are significantly different from those for severe cases of MERS-CoV but not for mild cases of MERS-CoV.

**Table 1.** Estimated parameters for SARS-CoV-2 and MERS-CoV infection obtained from throat swabs

| Parameter Name | Symbol (Unit) | SARS-CoV-2 | MERS-CoV |  |
| --- | --- | --- | --- | --- |
|  |  |  | Mild | Severe |
| Parameters obtained from fitting to the clinical time-series datasets |  |  |  |  |
| Maximum rate constant for viral replication | $\gamma$ (day<sup>-1</sup>) | 4.55<sup>†</sup> | 3.36 | 3.76 |
| Rate constant for virus infection | $\beta$ ((copies/ml)<sup>-1</sup> day<sup>-1</sup>) | $6.77 \times 10^{-5}$ | $2.10 \times 10^{-6}$ | $1.10 \times 10^{-6}$ |
| Death rate of infected cells | $\delta$ (day<sup>-1</sup>) | 1.59 | 2.47 | 1.32 |
| Viral load at symptom onset | $V(0)$ (copies/ml) | 21.8 | $2.43 \times 10^{-4}$ | $1.21 \times 10^{-3}$ |
| Quantities derived from estimated parameters |  |  |  |  |
| Within-hots basic reproduction number | $R_0$ | 2.87 | 1.36 | 2.84 |
| Critical inhibition rate | $C^*$ | 0.65 | 0.26 | 0.65 |
| Length of virus production | $L$ | 0.63 | 0.41 | 0.76 |
| Length of viral load above detection limit | $T_{VL}$ (days) | 9.34 | 10.9 | 14.0 |

<sup>†</sup> Median value

**Table 2.** Estimated parameters for SARS-CoV and MERS-CoV infection obtained from nasopharyngeal/sputum/tracheal aspirate

| Parameter Name | Symbol (Unit) | SARS-CoV | MERS-CoV |  |
| --- | --- | --- | --- | --- |
|  |  |  | Mild | Severe |
| Parameters obtained from fitting to the clinical time-series datasets |  |  |  |  |
| Maximum rate constant for viral replication | $\gamma$ (day<sup>-1</sup>) | 3.09<sup>†</sup> | 3.14 | 3.10 |
| Rate constant for virus infection | $\beta$ ((copies/ml)<sup>-1</sup> day<sup>-1</sup>) | $8.24 \times 10^{-3}$ | $8.37 \times 10^{-2}$ | $1.31 \times 10^{-3}$ |
| Death rate of infected cells | $\delta$ (day<sup>-1</sup>) | 0.66 | 0.67 | 0.46 |
| Viral load at symptom onset | $V(0)$ (copies/ml) | 2.54 | 5.89 | 3.05 |
| Quantities derived from estimated parameters |  |  |  |  |
| Within-host basic reproduction number | $R_0$ | 4.67 | 4.66 | 6.72 |
| Critical inhibition rate | $C^*$ | 0.79 | 0.79 | 0.85 |
| Length of virus production | $L$ | 1.51 | 1.49 | 2.17 |
| Length of viral load above detection limit | $T_{VL}$ (days) | 29.6 | 31.9 | 16.3 |

<sup>†</sup> Median value

### Characterising SARS-CoV and MERS-CoV infections by analysing viral load measurements collected from nasopharyngeal/sputum/tracheal aspirate

To extend our analysis to include SARS-CoV, we analysed SARS-CoV viral loads in nasopharyngeal aspirate reported by Peiris *et al. (12)* and MERS-CoV viral loads reported by Oh *et al*. (8) in sputum or tracheal aspirate. The estimated parameters, viral load at symptom onset, and the indices derived from the estimated parameters are listed in **Table 1** and **Fig. 2**, and the typical behaviour of the model is shown together with the data in **Fig. 1B** for SARS-CoV (blue) and MERS-CoV (black or grey). The estimated values of *L* for SARS-CoV and MERS-CoV are not significantly different. Surprisingly, the estimated values of *R*_0_ and *C*^*∗*^ for SARS-CoV are significantly different from those for severe cases of MERS-CoV (*p* = 0.03 and 0.02 from bootstrap *t*-test), but not for mild MERS-CoV cases (*p* = 0.52 and 0.47 from bootstrap *t*-test) (**Fig. 2**). This demonstrates that *in vivo* viral dynamics of SARS-CoV infection are similar to those for MERS-CoV in mild cases but not in severe cases. Collectively, the findings from the viral load data analyses for the two different specimens (throat/nasal swabs and nasopharyngeal/sputum/tracheal aspirate) implied that SARS-CoV-2 also causes infection more effectively than SARS-CoV.

### Evaluation of anti-SARS-CoV-2 therapies

Our quantitative analyses provide insights into optimal usage of anti-SARS-CoV-2 therapies under development. In particular, it remains poorly understood how a delay of treatment initiation after primary infection, or how incomplete blocking of virus infection/replication, impacts the viral load dynamics. Based on our mathematical model and estimated parameter values (**Table 1**), we conducted *in silico* experiments for possible anti-SARS-CoV-2 therapies to investigate the expected outcome under hypothetical drug therapies (or vaccine use) possessing different antiviral mechanisms (**Fig. 3**).

**Fig. 3.**
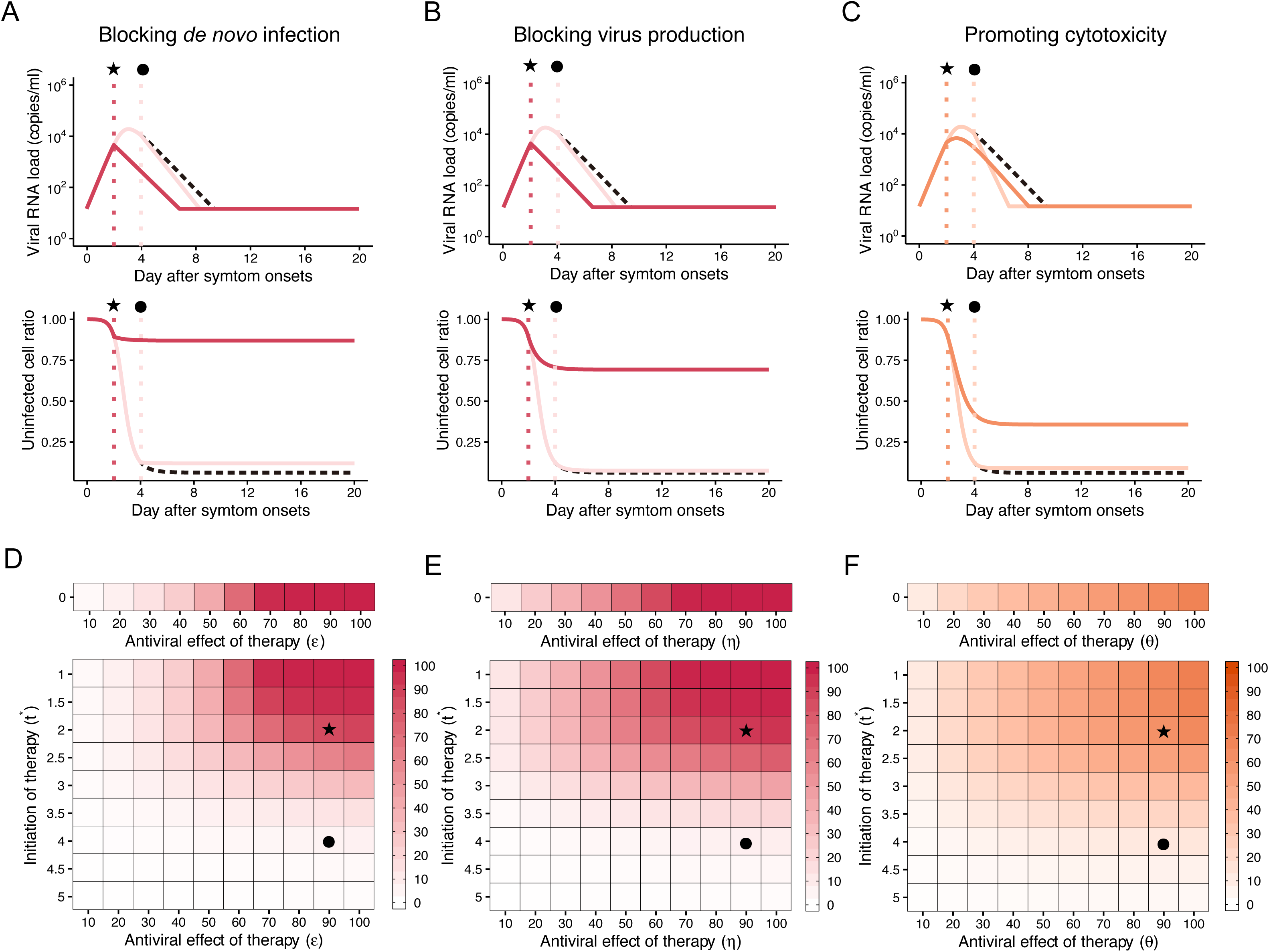
*In silico* experiments to predict the outcomes of possible anti-SARS-CoV-2 therapies. In each case, the therapy was initiated after 2 (★) or 4 (*•*) days from symptom onset with 90% inhibition rate. The expected dynamics of the viral loads (top) and the uninfected target cell ratio (bottom) under the hypothetical therapy (antiviral drug or vaccine) for blocking *de novo* infection, virus production, and promoting cytotoxic effects are shown in (**A**), (**B**) and (**C**), respectively, using the median values of our estimated parameters and an initial viral load that is equal to the detection limit. The coloured solid curves and black dashed curves correspond to SARS-CoV-2 infection dynamics with and without the therapies. In addition, we simulated the model for a range of therapy efficacies and times at which therapies were introduced, and the results are summarised in (**D**), (**E**) and (**F**) for therapies that block *de novo* infection, block virus production and promote cytotoxic effects, respectively. Darker regions of these panels indicate a larger reduction in AUC of viral load which implies a stronger antiviral effect against SARS-CoV-2 infection. The expected values with *t*^*^ =0 indicate the antiviral effect of pre-exposure vaccines (or post-exposure prophylactic use of antivirals).

#### (i) Blocking *de novo* infection

One of the major mechanisms of action for antivirals is blocking *de novo* infections. This can be induced by drugs including human neutralising antibodies, viral entry-inhibitors and/or antibodies raised by vaccination (13, 14). For example, a SARS-CoV-specific human monoclonal antibody has been reported to cross react with SARS-CoV-2 (14). We conducted *in silico* experiments with varying drug efficacy (considering inhibition rates from 10% to 100%, i.e. 0.1 ≤ *ε* ≤ 1) and with the timing of initiation of therapy from 0 days (i.e., post-exposure prophylactic use of antivirals) until 5 days after symptom onset (i.e., 0 ≤ *t*^*∗*^ ≤ 5) (see **Methods**). Our results show that early initiation of therapy (especially within two to three days) with even a relatively weak drug (inhibition rates as low as 50%) might effectively reduce the area under the curve of viral load (AUC) and prevent significant reductions in the numbers of target cells because of cytopathic effects due to cell invasion. A therapy of this type initiated four days after symptom onset, on the other hand, is not predicted to induce a clear antiviral effect (**Fig. 3AD**). This suggests that blocking *de novo* infections is not likely to be effective unless the intervention is initiated before the peak viral load. Hence, appropriate initiation timing (i.e., before or very soon after symptom onset) is an important factor for suppressing viral load in addition to the therapy having the potential for antiviral effects.

#### (ii) Blocking virus production

The majority of antiviral drugs inhibit intracellular virus replication. Although their antiviral efficacies need to be confirmed, lopinavir/ritonavir (HIV protease inhibitors), remdesivir (anti-Ebola virus disease candidate) and other nucleoside analogues, and interferon have the potential to suppress SARS-CoV-2 by blocking virus production(15, 16). Interestingly, our results suggest that, even for relatively small inhibition rates of around 30%, the AUC of viral load is partially reduced if therapy is initiated early (within three days after symptom onset) (**Fig. 3BE**). However, if treatment is applied after the peak viral load, even drugs with 100% inhibition rate are not able to reduce viral loads, which is similar to the predicted outcomes of *de novo* blocking therapy.

#### (iii) Promoting cytotoxicity

Another possible antiviral mechanism is cytotoxic effects by adaptive immunity including those mediated by cytotoxic T lymphocytes. Here, we assume that promoting cytotoxicity increases the virus death rate by at most two times (i.e.,≤ *θ* ≤ 1), that is, achieves up to 50% reduction of the mean length of virus production. Compared with the other two therapies (blocking *de novo* infection and virus production), the induction of cytotoxicity had relatively mild effects on the AUC reduction if initiated before peak viral load. However, cytotoxicity induction initiated after peak viral load could effectively reduce the viral load AUC (**Fig. 3CF**). This implies that there is an optimal time to apply this therapy, and that significant antiviral effects are expected unless the promoting rate is too low or therapy is initiated either too early or too late. However, large reductions of target cells due to ongoing *de novo* infection cannot be avoided even with very early initiation (i.e., immediately after symptom onset) of the therapy.

## Conclusions

There are a number of potential transmission routes of SARS-CoV-2, including direct person-to-person transmission due to viral particle inhalation and contact transmission due to contact with nasal/oral/eye mucuous membranes. The risk of transmission depends on the viral load of the potential infector. Consequently, treatments reducing the viral load are important for the prevention of secondary transmission and aid population-scale outbreak control. We characterised viral infection dynamics using a mathematical model, and assessed potential strategies to reduce viral loads. Our analyses showed that both blocking *de novo* infection and virus production effectively reduces AUC of SARS-CoV-2 load; for example, if the therapy can reduce more than 90% of *de novo* infections and is initiated 3 days after symptoms onset, the viral load AUC is expected to be reduced by 81.4% (**Fig. 3DE**). However, if therapy is initiated after peak viral load (more than 2-3 days following symptom onset), the effect on viral load AUC is limited. Compared with either blocking *de novo* infection or virus production, promoting cytotoxicity showed relatively mild effects on AUC reduction, however initiation of that therapy after the peak viral load has the potential to still reduce viral load AUC (**Fig. 3F**).

The effectiveness of the hypothetical drugs can be evaluated in detail using a cell culture system supporting SARS-CoV-2 infection (13). Wang *et al*. proposed different classes of drugs for treating SARS-CoV-2 infections: chloroquine inhibited viral entry and remdesivir suppressed the virus post-entry, likely by suppressing viral replication (13). Although animal models for testing treatments have not been reported for SARS-CoV-2, a number of animal models exist for SARS-CoV including mice, hamsters, ferrets, and macaques (17). Animals could be used to verify the conclusions from our models, by monitoring the viral loads in animals treated with different types of drugs at different doses and different initiation timings. Such viral load data would allow further investigation of the effectiveness of drugs with different action mechanisms, which would be informative for development of appropriate treatment strategies (i.e., the optimal dose/timing of antivirals) for SARS-CoV-2 infections.

In conclusion, effective treatment of SARS-CoV-2 infections requires an appropriate choice of class-specific drugs; otherwise, the antivirals do not alter the viral load significantly and are wasted. Identification of SARS-CoV-2-specific virus characteristics is needed to design optimal treatments and to ensure that limited resources are deployed effectively. Additionally, effective combinations of anti-SARS-CoV-2 drugs and vaccines will maximise the impacts of control, reduce the required drug dose and potentially limit side effects, all of which are highly desirable. Our theoretical approach could complement ongoing experimental investigations into SARS-CoV-2 infection in BSL-3 laboratories and help establish a basis for COVID-19 treatment. To our knowledge, previous studies have neither characterised SARS-CoV-2 dynamics in humans using viral dynamics models, nor compared the resulting dynamics against those of other coronaviruses (i.e., SARS-CoV and MERS-CoV). Our mathematical modelling approach has led to an improved understanding of the characteristics of SARS-CoV-2 *in vivo*, and can be used to test possible treatments for COVID-19 further going forwards.

## Methods

### Study data

The data examined in our manuscript came from studies of SARS-CoV-2, MERS-CoV and SARS-CoV by Zou *et al. (1)*, Oh *et al*. (8) and Peiris *et al. (12)*, respectively. To extract the data from images in those publications, we used the program datathief III (version 1.5, Bas Tummers, www.datathief.org). We excluded patients for whom data were measured on only one day, and assumed that viral load values under the detection limit were set as the detection limit for the purposes of fitting the model. We converted cycle threshold (Ct) values reported in Zou *et al. (1)*, Oh *et al*. (8) and Peiris *et al. (12)* to viral RNA copies number values, where these quantities are inversely proportional to each other (18).

### Mathematical model

To parameterise coronavirus infection dynamics from patient viral load data, we derived a simplified mathematical model from the following virus dynamics model:

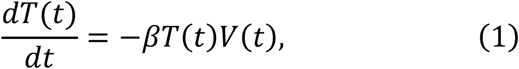

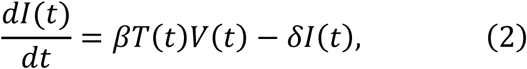

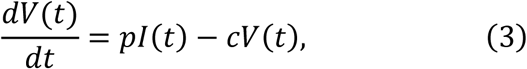

where the variables *T*(*t*), *I*(*t*) and *V*(*t*) are the numbers of uninfected target cells, infected target cells, and the amount of virus at time *t*, respectively. The parameters *β, δ, p*, and *c* represent the rate constant for virus infection, the death rate of infected cells, the viral production rate, and the clearance rate of the virus, respectively. Since the clearance rate of virus is typically much larger than the death rate of the infected cells *in vivo* (10, 19, 20), we made a quasi-steady state (QSS) assumption, *dV*(*t*)*⁄dt* = 0, and replaced Eq.(3) with *V*(*t*) = *pI*(*t*)*⁄c*. Because data on the numbers of coronavirus RNA copies, *V*(*t*), rather than the number of infected cells,*I*(*t*), were available, then *I* (*t*) = *cV*(*t*)*⁄p* was substituted into Eq.(2) to obtain

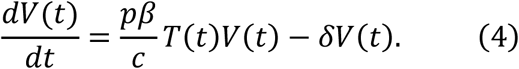

Furthermore, we defined the ratio of the number of uninfected target cells at time *t* to the initial number of uninfected target cells *T*(0), that is, *f*(*t*) = *T*(*t*)*⁄T*(0). Accordingly, we obtained the following simplified mathematical model, which we employed to analyse the data in this study:

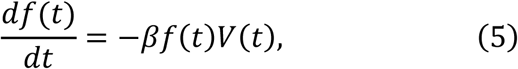

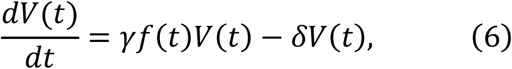

where *γ* = *pβ T*(0)/*c* is defined as the maximum viral replication rate for coronavirus infections. Note that the ratio *f*(*t*) is always less than or equal to 1.

In our analyses, the variable *V*(*t*) corresponds to the viral load in throat swabs for SARS-CoV-2 and MERS-CoV and in nasopharyngeal/sputum/tracheal aspirate for SARS-CoV and MERS-CoV. In the case of acute coronavirus infection, the loss of target cells by physiological turnover could be ignored, considering the long life-span of the target cells. Patient viral load data for SARS-CoV-2/SARS-CoV/MERS-CoV were fitted using a nonlinear mixed-effect modelling approach (described below), which uses the whole sample to estimate population parameters but also account for inter-individual variation.

### *In silico* experiments for possible anti-SARS-CoV-2 therapies

By utilising our novel mathematical model and the estimated parameter values, we investigated the antiviral effects of unlicensed but developing (promising) drugs with the following different mechanisms of action, depending on inhibition rates and timings of therapy initiation: (i) blocking *de novo* infection (e.g. via human neutralising antibodies, viral entry-inhibitors and antibody levels raised by vaccination (13, 14)); (ii) blocking virus production (such as lopinavir/ritonavir (HIV protease inhibitors), remdesivir (an anti-Ebola virus candidate) and other nucleoside analogues, and interferon (15, 16)); and (iii) promoting cytotoxicity (by adaptive immunity such as cytotoxic T lymphocytes). To simulate possible variations in the viral load and in the target cell numbers under these different types of anti-SARS-CoV-2 therapy, the median parameter sets were used to predict the expected outcome of each therapy. In other words, even though no drug administration trials have been conducted yet, we were able to infer the efficacy of each drug treatment based on our *in silico* experiments. We implemented the different mechanisms of action in the model as follows:

#### (i) Blocking *de novo* infection

The antiviral effect of blocking *de novo* infection therapy (0 < *ε* ≤ 1. *ε* = 1 implies *de novo* infection is 100% inhibited) initiated at *t*^*∗*^ days after symptom onset was modelled by assuming:

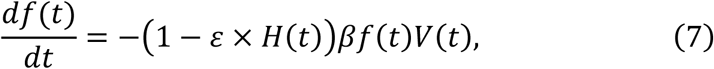

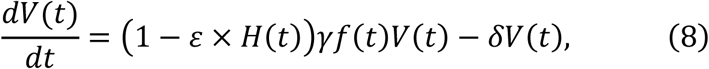

where *H*(*t*) is a Heaviside step function defined as *H*(*t*) = 0 if *t* < *t*^*∗*^ : otherwise *H*(*t*) = 1. We evaluated the expected antiviral effect of the therapy under different inhibition rates (*ε*) and initiation timings (*t*^*∗*^) using our estimated parameter values.

The mean reduction of cumulative virus production, i.e., the area under the curve of viral load (AUC: 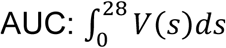 : because the observed durations COVID-19 infection are longer than previous coronavirus infection (i.e., SARS-CoV and MERS-CoV), we used the maximum length of observations 28 days as the upper bound for integration), induced by blocking *de novo* infection for SARS-CoV-2 was calculated. Note that the expected values at day 0 after symptom onset (*t*^*∗*^ = 0) corresponds to the antiviral effect of therapy initiated immediately after symptom onset.

#### (ii) Blocking virus production

Alternatively, we assumed an inhibition rate of virus production of 0 < *η* ≤ 1. The antiviral effect by blocking virus production (0 <*η* ≤ 1. *η* = 1 indicates that the virus reproduction from the infected cells are perfectly inhibited) is modelled as follows:

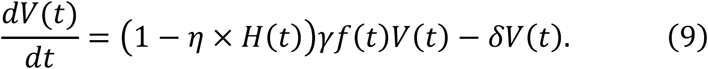

Note that the difference between blocking *de novo* infection and virus production is that the former reduces *β*, whereas the latter reduces*p* in the full model (1)-(3).

#### (iii) Promoting cytotoxicity

The antiviral effect of promoting cytotoxicity therapy (0 < *θ* ≤ 1. *θ* = 1 indicates that the mean duration of virus production is doubled) was modelled as follows:

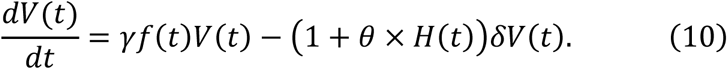

### The nonlinear mixed effect model

MONOLIX 2019R2 (www.lixoft.com), a program that implements a maximum likelihood estimation procedure for parameters in a nonlinear mixed-effects model, was employed to fit to the viral load data. Nonlinear mixed-effects modelling approaches allow a fixed effect as well as a random effect describing the inter-patient variability. Including a random effect amounts to a partial pooling of the data between individuals to improve estimates of the parameters applicable across the population of patients. By using this approach, the differences between viral dynamics in different patients were not estimated explicitly, nor did we fully pool the data which would bias estimates towards highly sampled patients. In this method of estimation, each parameter estimate 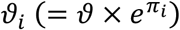 depends on the individual where *ϑ* is fixed effect, and π_*i*_ is random effect with an assumed Gaussian distribution with mean 0 and standard deviation Ω. Population parameters and individual parameters were estimated using the stochastic approximation expectation-maximisation algorithm and empirical Bayes’ method, respectively. Individual estimated parameters and initial values for patients are summarized in **Table S1**. Using estimated individual parameters and a Markov chain Monte Carlo algorithm, the conditional distribution is obtained which can represent the uncertainty in individual parameter values. We obtained 100 sets of estimated parameters for each individual patient by fitting the simplified mathematical model (Eqs. (5–6)) to the data. The estimation was performed for viral loads in throat swab from SARS-CoV-2, mild MERS-CoV, and severe MERS-CoV separately, and the distributions of the parameters were compared and tested using the bootstrap *t*-test. Due to the small sample size for viral loads in sputum/tracheal aspirate for MERS-CoV, we assumed the fixed effect was the same for mild and severe MERS-CoV cases. Otherwise, the process was exactly the same as that described for the throat swab data.

### The computation of *L, R*_0_, *C*^*∗*^ and *T*_*VL*_

Based on the estimated parameter distributions, we calculated several quantities: the duration of virus production (*L*), the basic reproduction number (*R*_0_), and the critical inhibition rate (*C*^*∗*^). We also calculated the period during which the viral load was above the detection limit (*T*_*VL*_) from the *in silico* simulations with individual estimated parameters and an initial viral load equal the detection limit (i.e. numerical experiments began at the point at which the virus became detectable). The distributional estimates of *R*_0_, *L, C*^*∗*^ and *T*_*VL*_ were calculated separately for SARS-CoV-2 and SARS-CoV, as well as for severe and mild cases of MERS-CoV.

## Data Availability

The data used in this study were extracted from the following papers:
1. Zou L, et al. (2020) SARS-CoV-2 Viral Load in Upper Respiratory Specimens of Infected Patients. N Engl J Med.
2. Oh MD, et al. (2016) Viral Load Kinetics of MERS Coronavirus Infection. N Engl J Med 375(13):1303-1305.
3. Peiris JS, et al. (2003) Clinical progression and viral load in a community outbreak of coronavirus-associated SARS pneumonia: a prospective study. Lancet 361(9371):1767-1772.

## Acknowledgments

This study was supported in part by Basic Science Research Program through the National Research Foundation of Korea funded by the Ministry of Education 2019R1A6A3A12031316 (to K.S.K.); Grants-in-Aid for JSPS Scientific Research (KAKENHI) Scientific Research B 17H04085 (to K.W.), 18KT0018 (to S.I.), 18H01139 (to S.I.), 16H04845 (to S.I.), Scientific Research S 15H05707 (to S.N.), Scientific Research in Innovative Areas 19H04839 (to S.I.), 18H05103 (to S.I.); AMED CREST 19gm1310002 (to S.I.); AMED J-PRIDE 19fm0208019j0003 (to K.W.), 19fm0208006s0103 (to S.I.), 19fm0208014h0003 (to S.I.), 19fm0208019h0103 (to S.I.); AMED Research Program on HIV/AIDS 19fk0410023s0101 (to S.I.); Research Program on Emerging and Re-emerging Infectious Diseases 19fk0108050h0003 (to S.I.); Program for Basic and Clinical Research on Hepatitis 19fk0210036j0002 (to K.W.), 19fk0210036h0502 (to S.I.); Program on the Innovative Development and the Application of New Drugs for Hepatitis B 19fk0310114j0003 (to K.W.), 19fk0310101j1003 (to K.W.), 19fk0310103j0203 (to K.W.), 19fk0310114h0103 (to S.I.); JST PRESTO (to S.N.); JST MIRAI (to K.W. and S.I.); JST CREST (to K.W. and S.I.); The Yasuda Medical Foundation (to K.W.); Smoking Research Foundation (to K.W.); Takeda Science Foundation (to K.W.); Mochida Memorial Foundation for Medical and Pharmaceutical Research (to K.W.); Mitsui Life Social Welfare Foundation (to S.I. and K.W.); Shin-Nihon of Advanced Medical Research (to S.I.); Suzuken Memorial Foundation (to S.I.); Life Science Foundation of Japan (to S.I.); SECOM Science and Technology Foundation (to S.I.); The Japan Prize Foundation (to S.I.); Toyota Physical and Chemical Research Institute (to S.I.); Fukuoka Financial Group, Inc. (to S.I.); Kyusyu Industrial Advancement Center Gapfund Program (to S.I.); Foundation for the Fusion Of Science and Technology (to S.I.); a Junior Research Fellowship from Christ Church, Oxford (to R.N.T.)

## Competing interests

The authors declare that they have no competing interests.

## Authors’ contributions

Conceived and designed the study: KE KW RNT SI. Analysed the data: KSK KE YI SI HO YK SN SI. Wrote the paper: KSK KE KW RNT SI. All authors read and approved the final manuscript.

## Supplementary Appendix

**Table S1.** Individual estimated parameters and initial values

| Patient ID | $\gamma$ | $\beta$ | $\delta$ | $V(0)$ | $L$ | $R_0$ | $C^*$ | $T_{VL}$ |
| --- | --- | --- | --- | --- | --- | --- | --- | --- |
| SAVS-CoV-2 patients: datasets are obtained from throat swabs |  |  |  |  |  |  |  |  |
| C | 4.85 | $2.08 \times 10^{-4}$ | 1.64 | $1.66 \times 10^3$ | 0.61 | 2.96 | 0.66 | 7.7 |
| D | 4.7 | $2.42 \times 10^{-5}$ | 1.52 | $3.71 \times 10^5$ | 0.66 | 3.09 | 0.68 | 10.4 |
| E | 3.65 | $1.51 \times 10^{-5}$ | 1.7 | $2.11 \times 10^{-5}$ | 0.59 | 2.15 | 0.53 | 13.2 |
| F | 4.86 | $4.15 \times 10^{-5}$ | 1.61 | 34.9 | 0.62 | 3.02 | 0.67 | 9.4 |
| H | 4.7 | $5.92 \times 10^{-4}$ | 1.77 | $6.1 \times 10^3$ | 0.56 | 2.66 | 0.62 | 6.6 |
| I | 4.47 | $8.08 \times 10^{-8}$ | 1.46 | 1.18 | 0.68 | 3.06 | 0.67 | 17.4 |
| K | 4.7 | $3.97 \times 10^{-5}$ | 1.58 | 9.1 | 0.63 | 2.97 | 0.66 | 9.7 |
| L | 3.89 | $6.55 \times 10^{-6}$ | 1.65 | $0.14 \times 10^{-2}$ | 0.61 | 2.36 | 0.58 | 13.5 |
| N | 4.7 | $3.97 \times 10^{-5}$ | 1.58 | 9.1 | 0.63 | 2.97 | 0.66 | 9.7 |
| O | 4.86 | $1.09 \times 10^{-4}$ | 1.63 | 26.7 | 0.61 | 2.98 | 0.66 | 8.4 |
| P | 4.7 | $3.97 \times 10^{-5}$ | 1.58 | 9.1 | 0.63 | 2.97 | 0.66 | 9.7 |
| Q | 4.63 | $2.29 \times 10^{-5}$ | 1.60 | 8.38 | 0.63 | 2.89 | 0.65 | 10.4 |
| S | 4.7 | $3.97 \times 10^{-5}$ | 1.58 | 9.1 | 0.63 | 2.97 | 0.66 | 9.7 |
| T | 4.49 | $5.48 \times 10^{-5}$ | 1.57 | 2.27 | 0.64 | 2.86 | 0.65 | 9.7 |
| MERS-CoV severe patients: datasets are obtained from throat swabs |  |  |  |  |  |  |  |  |
| 1 | 3.57 | $2.61 \times 10^{-7}$ | 1.16 | $0.80 \times 10^{-3}$ | 0.86 | 3.08 | 0.68 | 13.7 |
| 2 | 3.7 | $1.00 \times 10^{-6}$ | 1.44 | $0.99 \times 10^{-3}$ | 0.69 | 2.57 | 0.61 | 11.0 |
| 3 | 3.82 | $2.21 \times 10^{-6}$ | 1.57 | $2.08 \times 10^{-3}$ | 0.64 | 2.43 | 0.59 | 9.6 |
| 4 | 3.00 | $1.00 \times 10^{-6}$ | 1.48 | $0.33 \times 10^{-3}$ | 0.68 | 2.03 | 0.51 | 13.2 |
| 5 | 3.69 | $9.96 \times 10^{-6}$ | 1.43 | $1.03 \times 10^{-3}$ | 0.70 | 2.58 | 0.61 | 11.0 |
| 6 | 3.64 | $1.43 \times 10^{-6}$ | 1.54 | $1.19 \times 10^{-3}$ | 0.65 | 2.36 | 0.58 | 10.6 |
| 7 | 4.09 | $1.97 \times 10^{-6}$ | 1.19 | $2.79 \times 10^{-2}$ | 0.84 | 3.44 | 0.71 | 9.9 |
| 8 | 3.38 | $1.34 \times 10^{-6}$ | 2.3 | $0.47 \times 10^{-3}$ | 0.43 | 1.47 | 0.32 | 13.6 |
| 9 | 4.2 | $2.04 \times 10^{-6}$ | 1.58 | $4.22 \times 10^{-3}$ | 0.63 | 2.66 | 0.62 | 9.0 |
| MERS-CoV mild patients: datasets are obtained from throat swabs |  |  |  |  |  |  |  |  |
| 10 | 3.35 | $1.46 \times 10^{-6}$ | 2.63 | $0.60 \times 10^{-3}$ | 0.38 | 1.27 | 0.21 | 16.3 |
| 11 | 3.36 | $1.46 \times 10^{-6}$ | 2.46 | $0.58 \times 10^{-3}$ | 0.41 | 1.37 | 0.27 | 14.6 |
| 12 | 3.7 | $2.61 \times 10^{-6}$ | 1.97 | $0.49 \times 10^{-3}$ | 0.51 | 1.88 | 0.47 | 9.8 |
| 13 | 4.19 | $3.84 \times 10^{-6}$ | 1.98 | $0.36 \times 10^{-3}$ | 0.51 | 2.12 | 0.53 | 8.1 |
| 14 | 3.36 | $1.46 \times 10^{-6}$ | 2.53 | $0.63 \times 10^{-3}$ | 0.40 | 1.33 | 0.25 | 15.2 |
| 15 | 3.36 | $1.46 \times 10^{-6}$ | 2.46 | $0.58 \times 10^{-3}$ | 0.41 | 1.37 | 0.27 | 14.6 |
| 16 | 3.52 | $1.46 \times 10^{-6}$ | 1.76 | $0.81 \times 10^{-3}$ | 0.57 | 2.00 | 0.50 | 11.0 |
| 17 | 3.47 | $1.46 \times 10^{-6}$ | 1.89 | $0.68 \times 10^{-3}$ | 0.53 | 1.84 | 0.46 | 11.3 |
| SAVS-CoV patients: datasets are obtained from nasopharyngeal aspirate |  |  |  |  |  |  |  |  |
| A | 3.05 | $2.59 \times 10^{-8}$ | 0.70 | 1.87 | 1.43 | 4.36 | 0.77 | 23.0 |
| B | 3.07 | $7.67 \times 10^{-8}$ | 0.70 | 2.28 | 1.43 | 4.39 | 0.77 | 20.6 |
| C | 3.12 | $2.41 \times 10^{-7}$ | 0.68 | 3.61 | 1.47 | 4.59 | 0.78 | 18.7 |
| D | 3.14 | $3.31 \times 10^{-8}$ | 0.52 | 4.55 | 1.92 | 6.04 | 0.83 | 26.7 |
| E | 3.05 | $1.51 \times 10^{-7}$ | 0.86 | 1.83 | 1.16 | 3.55 | 0.72 | 17.6 |
| F | 3.08 | $1.78 \times 10^{-7}$ | 0.82 | 2.37 | 1.22 | 3.76 | 0.73 | 17.6 |
| G | 3.06 | $6.67 \times 10^{-8}$ | 0.90 | 1.90 | 1.11 | 3.40 | 0.71 | 18.6 |
| H | 3.05 | $3.41 \times 10^{-8}$ | 0.65 | 1.82 | 1.54 | 4.69 | 0.79 | 23.4 |
| I | 3.03 | $2.82 \times 10^{-7}$ | 1.01 | 1.45 | 0.99 | 3.00 | 0.67 | 15.6 |
| J | 3.12 | $1.72 \times 10^{-8}$ | 0.51 | 3.97 | 1.96 | 6.12 | 0.84 | 28.8 |
| K | 3.11 | $2.62 \times 10^{-8}$ | 0.63 | 3.23 | 1.59 | 4.94 | 0.80 | 24.2 |
| L | 3.11 | $1.57 \times 10^{-7}$ | 0.65 | 3.31 | 1.54 | 4.78 | 0.79 | 20.0 |
| M | 3.13 | $3.22 \times 10^{-7}$ | 0.63 | 4.32 | 1.59 | 4.97 | 0.80 | 18.9 |
| N | 3.12 | $1.58 \times 10^{-8}$ | 0.55 | 3.80 | 1.82 | 5.67 | 0.82 | 27.6 |
| MERS-CoV severe patients: datasets are obtained from sputum or tracheal aspirate |  |  |  |  |  |  |  |  |
| 1 | 3.10 | $3.26 \times 10^{-8}$ | 0.40 | 3.14 | 2.50 | 7.75 | 0.87 | 39.5 |
| 2 | 3.12 | $3.79 \times 10^{-8}$ | 0.44 | 3.75 | 2.27 | 7.09 | 0.86 | 30.4 |
| 3 | 3.08 | $0.46 \times 10^{-8}$ | 0.47 | 2.56 | 2.13 | 6.55 | 0.85 | 33.9 |
| 4 | 3.09 | $1.16 \times 10^{-8}$ | 0.59 | 2.69 | 1.69 | 5.24 | 0.81 | 27.0 |
| 5 | 3.09 | $1.13 \times 10^{-8}$ | 0.42 | 2.75 | 2.38 | 7.36 | 0.86 | 34.7 |
| 6 | 3.10 | $5.84 \times 10^{-8}$ | 0.53 | 2.89 | 1.89 | 5.85 | 0.83 | 25.4 |
| MERS-CoV mild patient: datasets are obtained from sputum or tracheal aspirate |  |  |  |  |  |  |  |  |
| 10 | 3.16 | $5.84 \times 10^{-7}$ | 0.65 | 6.19 | 1.54 | 4.86 | 0.79 | 18.5 |
| 11 | 3.13 | $2.60 \times 10^{-6}$ | 0.62 | 4.85 | 1.61 | 5.05 | 0.80 | 14.8 |
| 12 | 3.12 | $4.63 \times 10^{-7}$ | 0.76 | 3.52 | 1.32 | 4.11 | 0.76 | 16.3 |

## Notes

### Competing Interest Statement

The authors have declared no competing interest.

## References

1. Zou L, et al. (2020) SARS-CoV-2 Viral Load in Upper Respiratory Specimens of Infected Patients. N Engl J Med.

2. Fry AM, et al. (2014) Efficacy of oseltamivir treatment started within 5 days of symptom onset to reduce influenza illness duration and virus shedding in an urban setting in Bangladesh: a randomised placebo-controlled trial. Lancet Infect Dis 14(2):109–118.

3. Li Q, et al. (2020) Early Transmission Dynamics in Wuhan, China, of Novel Coronavirus-Infected Pneumonia. N Engl J Med.

4. Thompson RN (2020) Novel Coronavirus Outbreak in Wuhan, China, 2020: Intense Surveillance Is Vital for Preventing Sustained Transmission in New Locations. J Clin Med 9(2).

5. Chan JF, et al. (2020) A familial cluster of pneumonia associated with the 2019 novel coronavirus indicating person-to-person transmission: a study of a family cluster. Lancet 395(10223):514–523.

6. Zhu N, et al. (2020) A Novel Coronavirus from Patients with Pneumonia in China, 2019. N Engl J Med 382(8):727–733.

7. Lu R, et al. (2020) Genomic characterisation and epidemiology of 2019 novel coronavirus: implications for virus origins and receptor binding. Lancet 395(10224):565–574.

8. Oh MD, et al. (2016) Viral Load Kinetics of MERS Coronavirus Infection. N Engl J Med 375(13):1303–1305.

9. Perelson AS (2002) Modelling viral and immune system dynamics. Nat Rev Immunol 2(1):28–36.

10. Martyushev A, Nakaoka S, Sato K, Noda T, & Iwami S (2016) Modelling Ebola virus dynamics: Implications for therapy. Antiviral Res 135:62–73.

11. Iwami S, et al. (2012) Identifying viral parameters from in vitro cell cultures. Front Microbiol 3:319.

12. Peiris JS, et al. (2003) Clinical progression and viral load in a community outbreak of coronavirus-associated SARS pneumonia: a prospective study. Lancet 361(9371):1767–1772.

13. Wang M, et al. (2020) Remdesivir and chloroquine effectively inhibit the recently emerged novel coronavirus (2019-nCoV) in vitro. Cell Res.

14. Tian X, et al. (2020) Potent binding of 2019 novel coronavirus spike protein by a SARS coronavirus-specific human monoclonal antibody. Emerg Microbes Infect 9(1):382–385.

15. Yao TT, Qian JD, Zhu WY, Wang Y, & Wang GQ (2020) A Systematic Review of Lopinavir Therapy for SARS Coronavirus and MERS Coronavirus-A Possible Reference for Coronavirus Disease-19 Treatment Option. J Med Virol.

16. Lu H (2020) Drug treatment options for the 2019-new coronavirus (2019-nCoV). Biosci Trends.

17. Peiris JS, Yuen KY, Osterhaus AD, & Stohr K (2003) The severe acute respiratory syndrome. N Engl J Med 349(25):2431–2441.

18. Poon LL, et al. (2004) Detection of SARS coronavirus in patients with severe acute respiratory syndrome by conventional and real-time quantitative reverse transcription- PCR assays. Clin Chem 50(1):67–72.

19. Ikeda H, et al. (2016) Quantifying the effect of Vpu on the promotion of HIV-1 replication in the humanized mouse model. Retrovirology 13:23.

20. Nowak MA & May RM (2000) Virus dynamics. (Oxford University Press Oxford).

